# MPN Transformation Is Characterized By Heterogeneous Shifts In Lineage Character Resulting In Both HSC-Like And More Differentiated Lineage Signatures

**DOI:** 10.1101/2024.04.16.24305909

**Authors:** Kamal Menghrajani, Deepika Dilip, Noushin Farnoud, Chris Famulare, Erin McGovern, Maria Sirenko, John Mascarenhas, Heidi Kosiorek, Ronald Hoffman, Ross Levine, Richard Koche, Raajit Rampal, Jacob Glass

## Abstract

Philadelphia chromosome-negative myeloproliferative neoplasms (MPNs) have a propensity to transform to an accelerated or blast phase (MPN-AP/BP). The resulting disease has clinically similar manifestations to Acute Myeloid Leukemia (AML) but worse clinical outcomes. Here we present the first comprehensive description of the transcriptomic characteristics of MPN-AP/BP. Our analysis incorporates data from 261 patients of the BeatAML cohort and 56 MPN-AP/BP patients, 11 of whom had paired samples from before and after transformation. We establish that transformed MPN is a transcriptionally distinct entity from de novo AML and chronic phase MPNs. Genomic pathways traditionally associated with MPN pathogenesis, such as IL2/STAT5 signaling, IL6/JAK/STAT3 signaling, and NUP98/HOXA9 fusions, were enriched in chronic-phase MPNs but are absent in transformed disease, suggesting JAK2 directed therapy may be less effective in this disease phase. We also discovered that gene expression signatures associated with doxorubicin resistance are highly enriched in transformed MPNs, which may explain the lack of efficacy of standard AML therapies. In addition, we identify that lineage composition at the time of transformation may define distinct subsets of MPN-AP/BP patients, which may assist in the future development of novel treatment strategies.

**Key Points:**

- Accelerated- and blast-phase transformed MPNs are a transcriptionally entities which are distinct from de novo AML.
- Transformed MPNs may be characterized by their lineage characteristics, which can drive clinical behavior and account for their inferior overall survival
- Gene expression signatures associated with doxorubicin resistance were highly enriched in transformed MPNs, which may explain the lack of effectiveness of anthracycline-based therapies

## Introduction

The Philadelphia chromosome-negative myeloproliferative neoplasms (MPNs) are characterized by the clonal proliferation of differentiated hematologic cells. Polycythemia vera (PV) manifests with increased production of red blood cells, essential thrombocythemia (ET) with increased platelets, and myelofibrosis (MF) with increased reticulin fibrosis in the bone marrow and frequent cytopenias^1,2^. MPNs are driven by hyperactive signaling through the JAK-STAT pathway, canonically triggered by the acquisition of somatic mutations in *JAK2, MPL*, or *CALR*^3,4^. While MPNs may remain stable for decades, the risk of transformation increases with disease duration. For patients with PV and ET, their disease may progress to secondary MF. Even more concerning is the potential for all MPNs to progress to accelerated-or blast-phase (MPN-AP/BP), which is clinically similar to acute myeloid leukemia (AML). Patients with MF have the highest risk of transformation – 10-20% at 10 years from diagnosis – while the risk among PV and ET patients is more modest, at 2-4% and 1-2%, respectively^5–10^.

Prior studies have evaluated the role of somatic mutations in the process of transformation, which has revealed that the spectrum of mutations in MPN-AP/BP appears distinct from that observed in de novo AML ^11^. However, the transcriptional alterations which underlie transformation to MPN-AP/BP, as well as how these alterations differ from those found in de novo AML, are poorly understood.

Specific analysis of the pathways involved in myeloid differentiation can aid in understanding how transformation alters the cellular composition of MPNs. Here, we use integrated RNAseq, genomic, cytogenetic, and clinical data from MPN-AP/BP patients and data from BeatAML to capture this transcriptional framework. We use a novel lineage deconvolution technique to quantify the proportion of each myeloid lineage stage present in each sample. We also include paired MPN samples from longitudinal follow-up of cases that allow for novel comparisons between the pre- and post-transformation states within single patients.

## Methods

### RNA Sequencing and Initial Data Processing

RNA-seq was performed on viably banked peripheral blood mononuclear cells for all MSK and MPN-RC samples at the MSK Integrated Genomics Operation and processed using the MSKCC Epigenetics Research and Innovation Lab computational pipeline. Briefly, 50bp paired end libraries were prepared and sequenced and the resulting FASTQ files were processed using trim_galore (version 0.6.4) for both adapter removal and read quality filtering with a quality threshold of 15. The processed FASTQ files were then aligned using STAR (version 2.7.10b) using default parameters in two pass mode. Post-alignment quality and coverage were assessed using the CollectRNASeqMetrics tool from Picard (version 2.18.6). Raw read counts were created using HTSeq (v0.9.1).

### Dataset Integration

Three different datasets were integrated for our analysis. The first dataset was from BeatAML, including chronic phase MPN, de-novo AML, and MPN AP/BP cases^12^. After excluding samples from patients with acute promyelocytic leukemia, RNA-seq data and clinical annotations were obtained for 281 patients, including both de-novo AML and MPN AP/BP cases. The second dataset included 45 patients from the MPN Research Consortium (MPN-RC) and Memorial Sloan Kettering Cancer Center (MSK) with MPN-AP/BP for whom both clinical data and tissue samples were available. The third dataset included paired chronic phase and MPN-AP/BP samples for patients seen at MSK, previously used to confirm LKB1/STK11 co-operativity seen in a murine MPN model^13^. This included eight patients who transformed from MPN to MPN-BP and three additional patients who transformed from chronic phase to MPN-AP. All of these specimens were pooled as post-transformation cases for purposes of the downstream analysis. For the MSK samples, we abstracted clinical data from the electronic medical record, ran genomic sequencing and RNAseq on patient samples, and performed downstream analyses through the Center for Epigenetics Research.

### Pathway Analysis: Transformed MPNs

The BeatAML and MPN-RC/MSK cohorts were batch corrected and processed through the DESeq2 software package, and differential expression analysis was run to compare transformed MPN (N = 76) and de-novo AML (N = 241) patients ^14^. Gene set enrichment analysis (GSEA) compareed pathways differing between AP/BP MPNs and de-novo AML cases. This was performed using the R fgsea package, with genes ranked according to the differential expression analysis^15^. Curated gene sets from MSigDB were queried using the msigdbr R package and filtered for biological relevance using key terms^16,17^. Gene set variation analysis (GSVA) was performed to generate a gene set enrichment score for each pathway in each patient and evaluate pathway variation between samples^18^. All AML samples from the BeatAML and MPN-RC/MSK cohorts were included to generate a background comparator for GSVA score generation, which allowed for a broader background for enrichment calculation. To separate samples based on variable regions, we filtered genes based on a standard deviation percentile of 90% and higher and applied hierarchical clustering (Ward’s method). To identify GSVA-based gene set enrichments in specific clusters a t-test was used to compare GSVA scores within the cluster(s) of interest to those in the remainder of the cohort. To identify enriched mutation frequency within a cluster, the Fisher’s exact test was employed.

### Pathway Analysis: High Risk MF

To further analyze early factors contributing to AML progression, the transcriptomic profiles of MF patients in the paired sample cohort who later developed MPN AP/BP were compared to an independent cohort of patients diagnosed with non-progressive chronic phase polycythemia vera (PV) over 5 years of observation. Differential expression analysis was performed in a similar fashion to the comparison of transformed MPN to de-novo AML. GSEA was similarly performed to identify differentially expressed pathways.

### Characterization of Lineage

Our custom RNA-seq lineage deconvolution algorithm quantified hematopoietic lineage in each sample and assessed changes between sample collection timepoints. RNA-seq data from normal hematopoietic stages were taken from the Corces et al. dataset and used to estimate hematopoietic stage proportions^19^. To integrate the BeatAML and MSK data with the Corces et al. batch, ComBat was applied for batch correction^20^. Thirteen hematopoietic stages were available to identify lineage among samples: HSC, MPP, LMPP, CMP, GMP, MEP, monocytes, erythroblast, CLP, CD4, CD8, B, and natural killer cells. To further characterize MPN aP/BP, we clustered these cases using the deconvolved lineage components and annotated cases with GSVA scores and mutation data.

### Survival Analysis

The lineage distribution of transformed MPN samples was analyzed using unsupervised hierarchical clustering. Kaplan-Meier curves were generated for each of the identified clusters to assess the impact of transformed MPN lineage subtype on survival. The transformed MPN and de novo AML cases were combined and stratified by risk for comparative survival analysis.

### *Pathway Analysis: MPN to* MPN AP/BP

Paired RNA-seq samples were batch corrected, normalized, and processed through the DESeq2 software package^14^. Differential expression analysis was performed between batch-corrected chronic phase MPN and de-novo AML?? MPN AP/BP samples while accounting for the patient of origin and a ranked gene list produced. This gene list was used to run a gene set enrichment analysis (GSEA) to compare both two groups. Among paired samples, a Wilcoxon signed-rank test was used to compare changes in hematopoietic stage proportion between the chronic phase MPN and transformed MPN time points.

## Results

To analyze the transcriptomic landscape of MPN AP/BP compared to de novo AML, data from 261 patients in the BeatAML cohort were integrated with 56 transformed MPN patients who had been seen and treated on MPN-RC clinical trials or at MSK^21,22^. Cases within the BeatAML cohort included those with transformed MPNs (n = 20), therapy-related AML (n = 41), and de novo AML (n = 200). Genomic sequencing data were available for 226 BeatAML patients. Of the 56 patients with MPN AP/BP from MSK, 11 had paired samples available from an earlier chronic phase timepoint (**Table 1**).

**Table 1:** Patient cohort composition. The patient cohort analyzed in this study is composed of a composite of patients on BeatAML and at our own institution (MSK). The MSK cohort included 11 paired samples, which are shown separately below.

|  | **<u>BeatAML</u>** | **<u>MSK</u>** | **<u>p</u>** |
| --- | --- | --- | --- |
| **<u>AML</u>** | **<u>261</u>** | **<u>56</u>** |  |
| Cytogenetics (%) |  |  | 0.003 |
| abn(17p) | 1 (0.4) | 1 (1.8) |  |
| CBF | 27 ( 10.3) | 0 (0.0) |  |
| Complex | 16 (6.1) | 5 (8.9) |  |
| del(5/7) | 6 (2.3) | 2 (3.6) |  |
| del20q | 1 (0.4) | 2 (3.6) |  |
| Monosomal | 26 ( 10.0) | 8 ( 14.3) |  |
| Normal | 108 ( 41.4) | 17 ( 30.4) |  |
| OND | 28 ( 10.7) | 4 (7.1) |  |
| t(3q26.2;v) | 3 (1.1) | 0 (0.0) |  |
| t(v;11q23) | 5 (1.9) | 0 (0.0) |  |
| t9.11 | 8 (3.1) | 0 (0.0) |  |
| tri8 | 7 (2.7) | 2 (3.6) |  |
| Unknown | 25 (9.6) | 15 ( 26.8) |  |
| PriorMPN = TRUE (%) | 20 (7.7) | 56 (100.0) | <0.001 |
| Relapsed.Residual = TRUE (%) | 10 (3.8) | 0 (0.0) | 0.286 |
| PriorMPNMoreThanTwoMths = TRUE (%) | 19 (7.3) | 26 ( 96.3) | <0.001 |
| Molecular.Data = TRUE (%) | 226 ( 86.6) | 55 ( 98.2) | 0.024 |
| tAML = TRUE (%) | 41 ( 15.7) | 0 (0.0) | 0.003 |
| **<u>MF</u>** | **<u>3\*</u>** | **<u>11</u>** |  |
| Cytogenetics (%) |  |  | 0.508 |
| abn(17p) | 0 (0.0) | 1 (9.1) |  |
| Complex | 0 (0.0) | 1 (9.1) |  |
| del(5/7) | 0 (0.0) | 1 (9.1) |  |
| del20q | 0 (0.0) | 2 ( 18.2) |  |
| Normal | 1 ( 33.3) | 4 ( 36.4) |  |
| OND | 1 ( 33.3) | 0 (0.0) |  |
| tri8 | 0 (0.0) | 1 (9.1) |  |
| Unknown | 1 ( 33.3) | 1 (9.1) |  |
| Molecular.Data = TRUE (%) | 3 (100.0) | 9 ( 81.8) | 1 |

\* Not used in paired transcriptomic analyses.

To compare MPNAP/BP to de novo and tAML, cases with both molecular and RNA-seq data were integrated across cohorts. These included 207 AML cases and 75 MPN-AP /BP cases (**Figure 1**). Unsupervised analysis was performed using hierarchical clustering applied to batch corrected, variance stabilized RNA-seq count data. After splitting the dataset into 6 clusters, clusters 4 and 5 are enriched for MPN AP/BP cases (N = 10, 33.3%; N = 36, 54.5%). Notably, the somatic mutation and cytogenetic profiles in these clusters were varied, suggesting that multiple genomic perturbations may lead to similar pathways of transformation from chronic phase MPNs toAP/BP. To assess higher-level perturbation of genomic pathways, gene set variation analysis (GSVA) was performed. This demonstrates the coalescence of transcriptomic perturbation into specific pathways that distinguish MPN-AP/BP from de novo AML cases.

**Figure 1.**
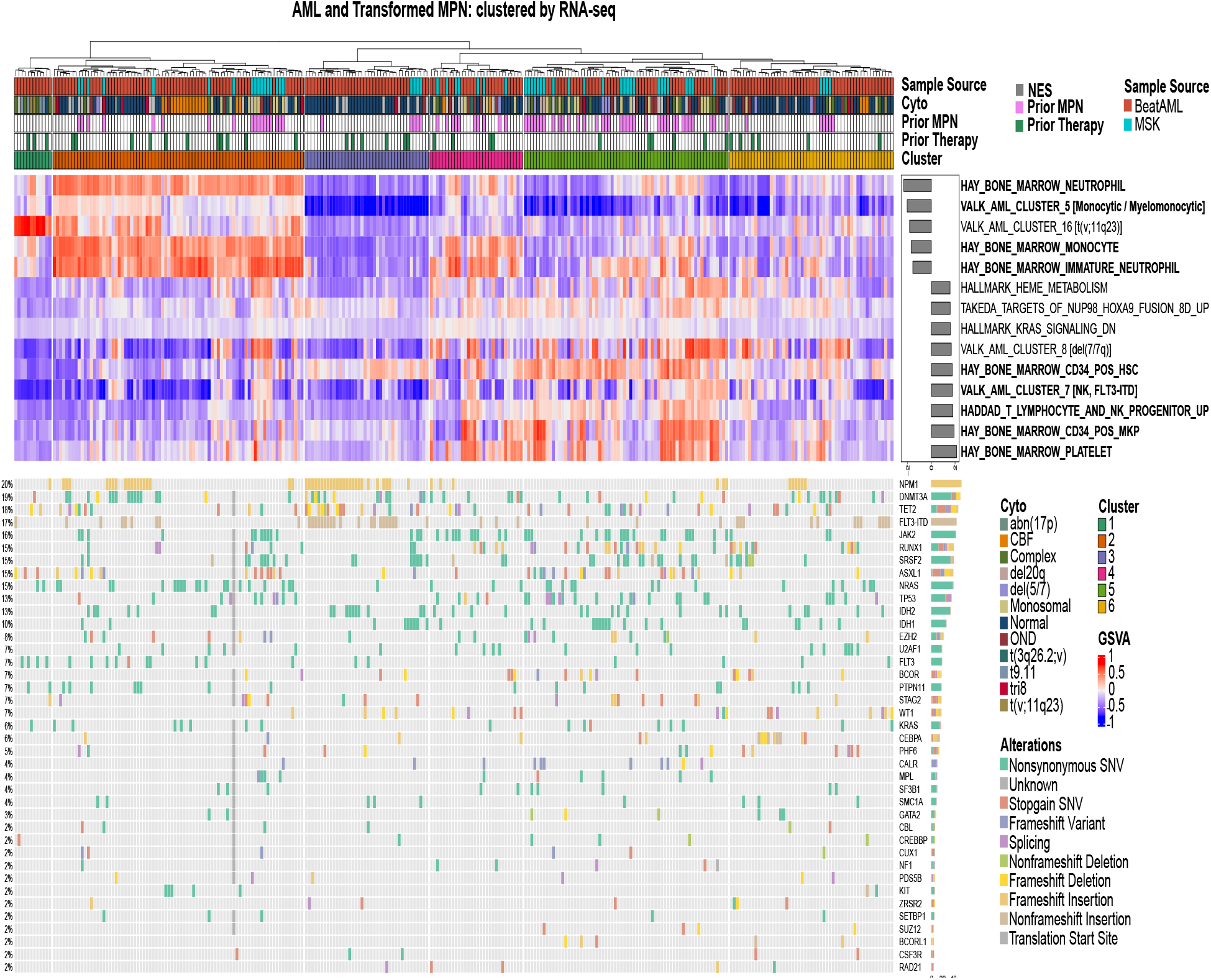
Integrated analysis of MSK and Beat AML cohorts. Following batch correction, hierarchical clustering of de novo AML and transformed MPN patient RNA-seq data was performed, revealing 6 distinct clusters. Gene set variation analysis (GSVA) of key differentially expressed pathways in the cohort is shown in heatmap format with normalized enrichment scores for post-MPN vs. other AML cases shown on the right. Molecular features, history of prior oncologic therapy (tAML), history or prior MPN, and batch are annotated at the top.

Interestingly, pathways enriched in cluster 5 and other MPN AP/BP consisted of hematopoietic stem cell gene sets (e.g. HAY_BONE_MARROW_CD34_POS_HSC: p < 0.01) while primary AML cases typically demonstrated upregulation in later stages of hematopoiesis, such as monocytes (HAY_BONE_MARROW_MONOCYTE: p < 0.01) as well as in previously identified AML-associated gene sets. Although less prominent than cluster 5, cluster 4 still had a significantly higher CD34-positive hematopoietic stem cell (HSC) GSVA score compared to other clusters (p < 0.001). This was driven in part by de-novo AML cases, suggesting that the stem-like phenotype is not exclusive to MPN AP/BP. Interestingly, cluster 2 and 4 show a general enrichment for immature neutrophil and monocyte gene sets compared to others (HAY_BONE_MARROW_IMMATURE_NEUTROPHIL: p<0.01, HAY_BONE_MARROW_MONOCYTE: p < 0.01). However cluster 2 diverged from cluster 4 in its strong enrichment for mature neutrophil character (HAY_BONE_MARROW_NEUTROPHIL p=0.02). Taken together, this would indicate a more differentiated phenotype than cluster 5, but less so than the de novo AML cases with mature neutrophil characteristics that dominate cluster 2. Cluster 1 is dominated by the previously described cluster 16 gene set described by Valk et al. Cluster 3 is partially enriched for the CD34+ HSC stage (HAY_BONE_MARROW_CD34_POS_HSC: p < 0.01) without enrichment for others. It is also enriched for NPM1 +/-FLT3-ITD mutations with subsets showing IDH1, IDH2, and IDH1/2 + SRSF2 mutation (Fisher’s exact: p<0.01). The several MPN-AP/BP cases in this cluster, marked by *JAK2* or *CALR* mutations, are likely similar to de-novo AML and tAML cases with a poorly differentiated phenotype.

In comparing the differential expression of specific genes between MPN-AP/BP and de novo / tAML, we found MIR29B1 was particularly upregulated in transformed MPNs (logFC = 9.52, q < 0.01), while the proto-oncogene CT45A1 had decreased expression in MPN-AP/BP compared to other AMLs (logFC = -7.47, q < 0.01) (**Figure 2A**). To evaluate specific gene sets differentially expressed between these two groups in a supervised fashion, gene set enrichment analysis (GSEA) was performed. This demonstrated that transformed MPNs were enriched for specific pathways characterizing earlier stages of differentiation including T-lymphocyte and NK cell progenitors and CD34-positive megakaryocyte progenitors (NES: 1.85, q<0.01; NES: 1.78, q=0.03). Interestingly, mature platelet pathways were also enriched (NES: 2.13, q < 0.01), which may reflect underlying MPN disease biology^23^.Pathways indicative of mature neutrophil character were enriched in the de novo / tAML AML cohort (NES: -1.57, q-value = 0.047) (**Figure 2B**).

**Figure 2.**
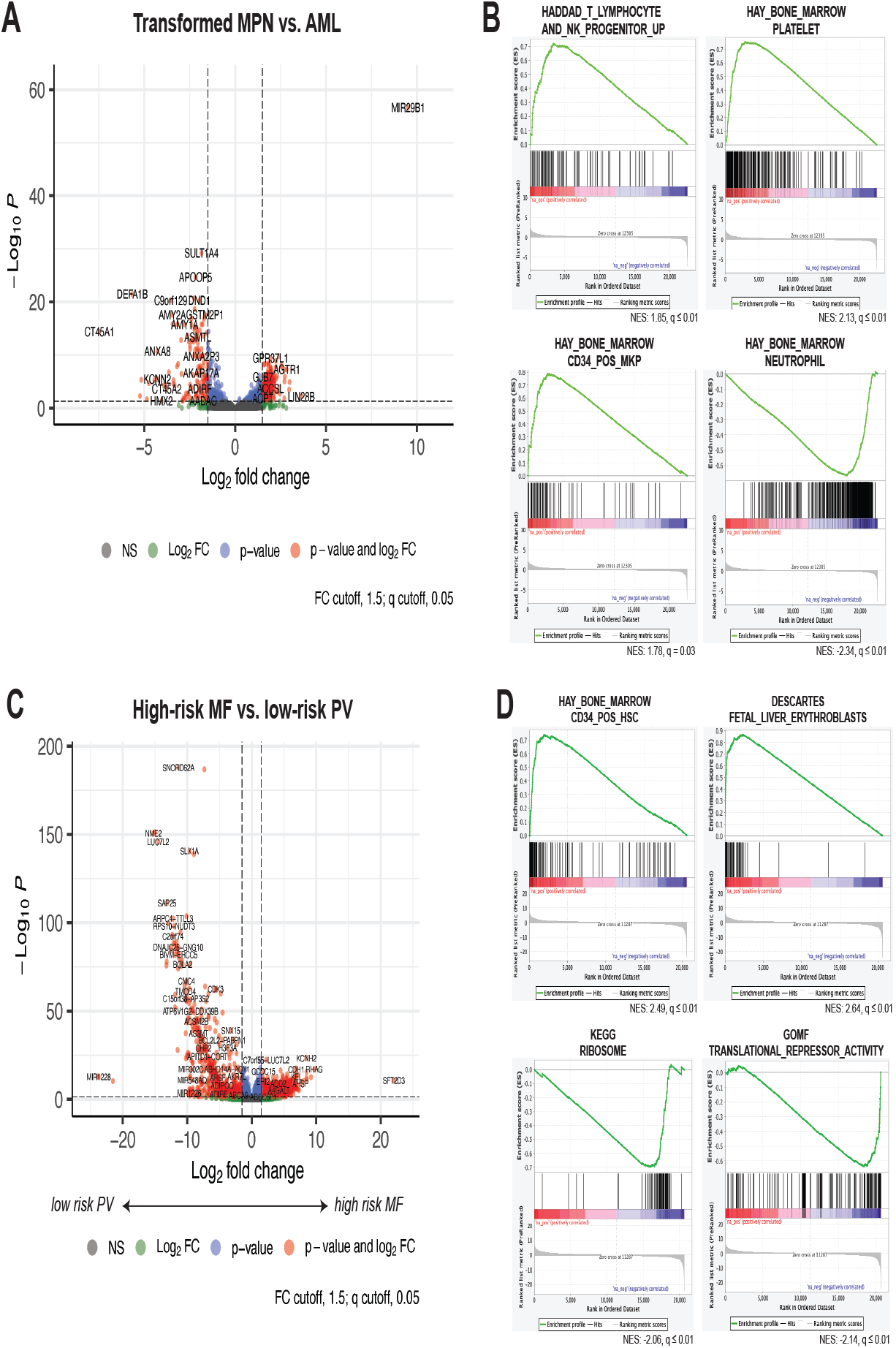
Gene expression and pathway analysis of transformed and high risk MPNs. A) Volcano plot comparing gene expression in blast phase MPN (MPN-BP) compared to other non-MPN associated AML. Log fold change gene expression is shown on the x-axis and -log_10_ adjusted p-value is shown on the y-axis. Colors denote log fold-change above 1.5 alone (green), corrected p-value < 0.05 (blue), neither (black), or both (red). B) Selected MSigDB GSEA enrichment plots for transformed MPN samples as compared to non-MPN associated AML. C) Volcano plot comparing gene expression in high-risk myelofibrosis (MF) patients compared to a cohort with low-risk polycythemia vera (PV). D) Selected MSigDB GSEA enrichment plots for High-risk MF as compared to low-risk PV.

We also compared MF cases with subsequent transformation to MPN-AP/BP to PV cases that did not show progression over a five-year observation period (**Figure 2C)**. We again observed enrichment of stem-like pathways in the high-risk cases including CD34-positive HSC (NES: 2.46, q-value = 3.66 x 10^-13^). We also noted downregulation of ribosomal activity (KEGG_RIBOSOME, NES = -2.07, q = 9.19 x 10^-6^) and upregulation of translational repression in high-risk MF cases (GOMF_TRANSLATIONAL_REPRESSOR_ACTIVITY, NES = -2.18, q = 1.32 x 10^-4^) (**Figure 2D**) . Two of the PV cases ultimately progressed to AML – one 6 years after initial evaluation, one to MF after 7 years and to AML after 8. Lineage deconvolution of these cases showed greater HSC and less MPP character than other PV cases in the cohort (**Figure S1**). Unsupervised analysis of the lineage deconvolution resulted in these samples clustering with MF cases rather than PV cases, suggesting the potential utility of lineage analysis as a prognostic tool. Interestingly, this was not reflected in the LSC-17 score, suggesting the specific utility of our deconvolution approach in MPNs compared to AML.

We next sought to characterize the specific transcriptomic changes defining the transformation of a chronic-phase MPN. Paired molecular and RNA-seq data were available from both the chronic-phase MPN (MF in all cases) and MPN AP/BP for 11 MSK patients. These data were used for a patient-adjusted differential expression analysis (**Figure 3, S2)**. Using a GSEA approach, we found again that the gene set for CD34-positive HSCs was enriched in the transformed MPNs relative to chronic phase (NES = 2.64, q-value < 0.0001). In addition, the gene set associated with E2F transcription factors was enriched (NES 2.66, q-value < 0.01), suggesting increased proliferation and DNA synthesis associated with transformation. The doxorubicin resistance gene set was also enriched (NES 2.40, q-value < 0.01), correlating with previously described clinical resistance to anthracycline-based chemotherapy transformed MPN patients^24,25^. Notably, gene sets associated with canonical pathways implicated in the pathogenesis of MPNs, such as IL2 / STAT5 signaling (NES -1.51, q = 0.002), IL6 / JAK / STAT3 signaling (NES -1.65, q = 0.0013), and NUP98 / HOXA9 fusions (NES -1.62, q = 0.0002), were all found to be enriched in the chronic-phase MPN compared to MPN AP/BP.

**Figure 3.**
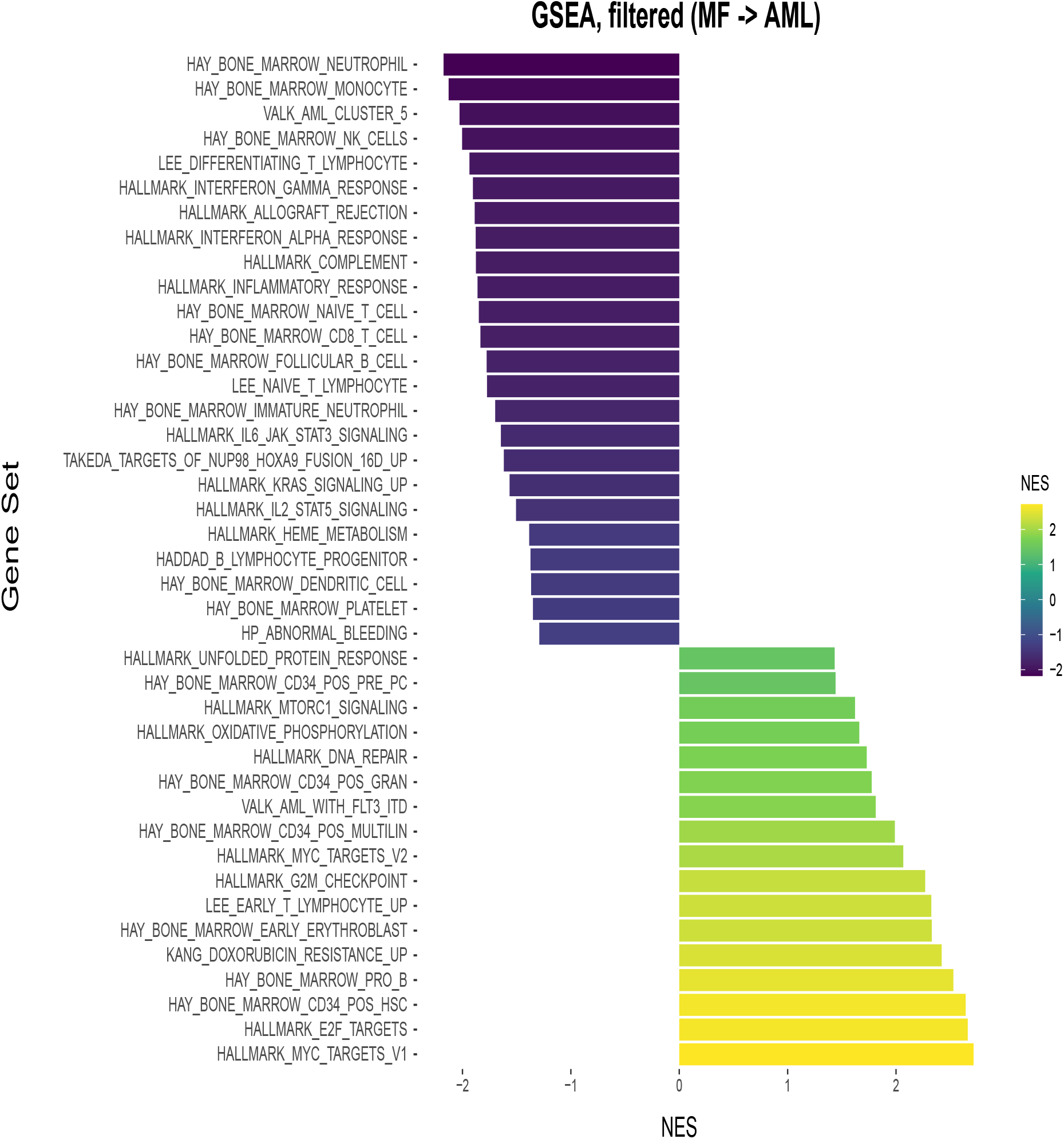
Gene set enrichment analysis of paired samples, comparing transformed disease to the prior myelofibrosis within the same patient. Normalized enrichment scores (NES) are shown. Gene sets for unrelated diseases and splitting up/down regulation were excluded as well as those with an absolute NES < 1.25 or with p-value > 0.05.

Given enrichment of the CD34-positive HSC gene set, we performed RNA-seq based lineage deconvolution on each of the paired samples to better understand sample-specific lineage shifts occurring with transformation (**Figure 4**). The time between chronic phase and BP samples varied from 3.2 months to 59.9 months. We found that MPNs transform in a heterogenous fashion reminiscent of the biological diversity in AML. In aggregate, transformation was associated with decreased monocytic (p = 0.002), natural killer (NK) (p = 0.042) character and expansion of common lymphoid progenitor characteristics (p = 0.022). Lineage composition changes varied, with certain samples showing HSC expansion (ID: 54910, ID: 46567) and others exhibiting LMPP expansion (ID: 29485, 2456). Interestingly, most samples showed a relative loss of monocyte character with transformation, potentially reflecting a shift from granulocyte production to other cell types, including myeloid-erythroid precursors (MEPs).

**Figure 4.**
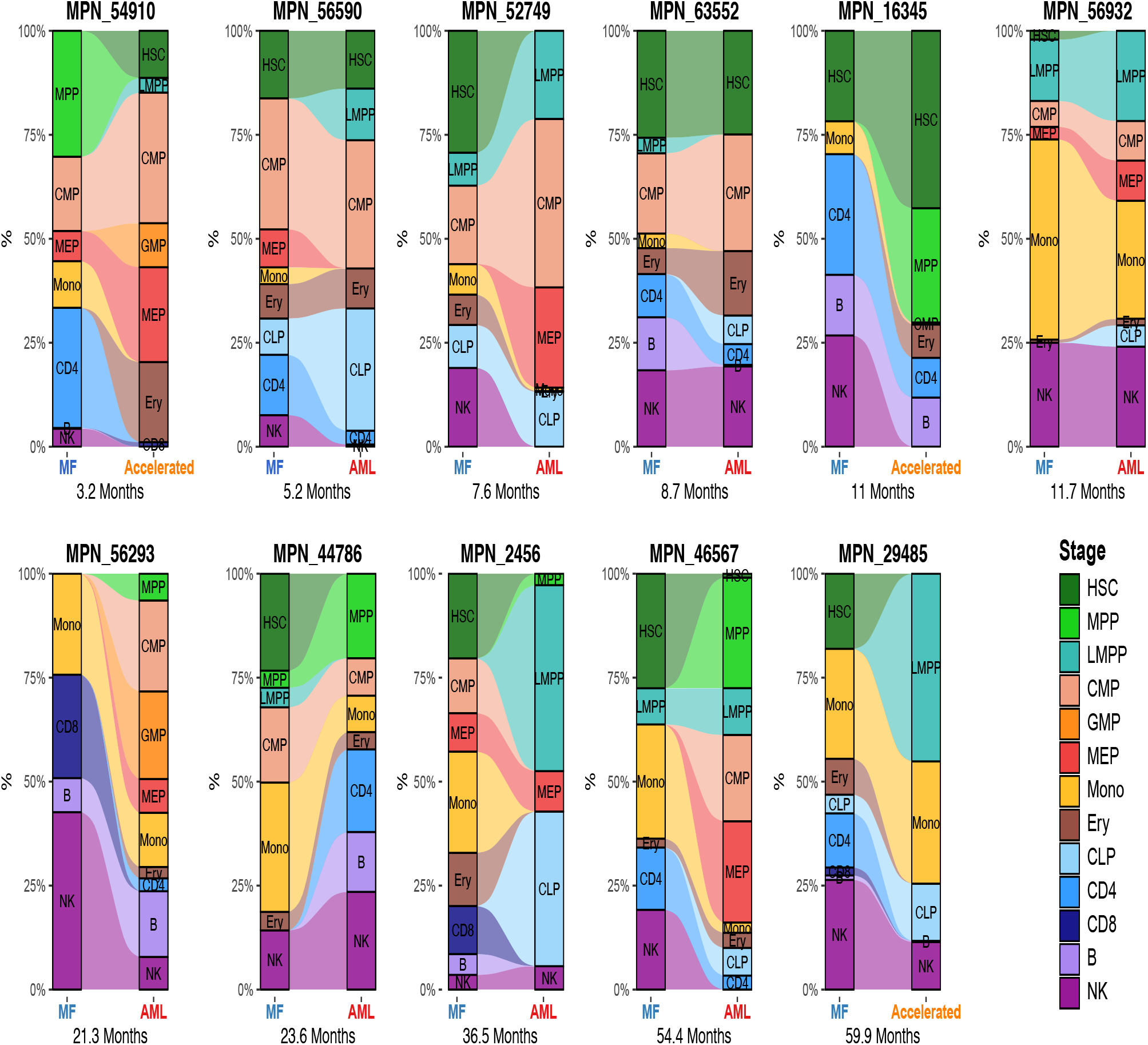
Comparison of lineage composition before and after MPN transformation. RNA-seq based lineage deconvolution was performed on each and illustrated as alluvial plots. Samples are ordered according to the time elapsing between the two sample collection dates.

To evaluate the patient specific lineage characteristics of MPN AP/BP in general, lineage deconvolution was performed on both the transformed MPN time point from MPN-RC/MSK samples and transformed MPNs in the BeatAML cohort (**Figure 5A**). Unsupervised analysis of the lineage composition of these cases showed several common lineage compositions within transformed MPNs. At the highest level of clustering, two groups emerged corresponding to the stage of differentiation. Clusters 1-3 show increased T cell, mono, and GMP character respectively while clusters 4-7 show increased MPP, LMPP, CMP, and HSC character. These roughly correspond to lower and higher GSVA scores for the CD34-positive HSC gene set. In addition, clusters 1-3 showed strong enrichment for immature neutrophil, neutrophil, and monocyte gene sets, consistent with a more differentiated character. Cluster size limits the ability to note molecular enrichments, however the general lack of consistent cytogenetic or molecular findings in each cluster suggests that transformed MPNs may be the result of convergent disease evolution from a diverse set of pathways from the chronic MPN state.

**Figure 5.**
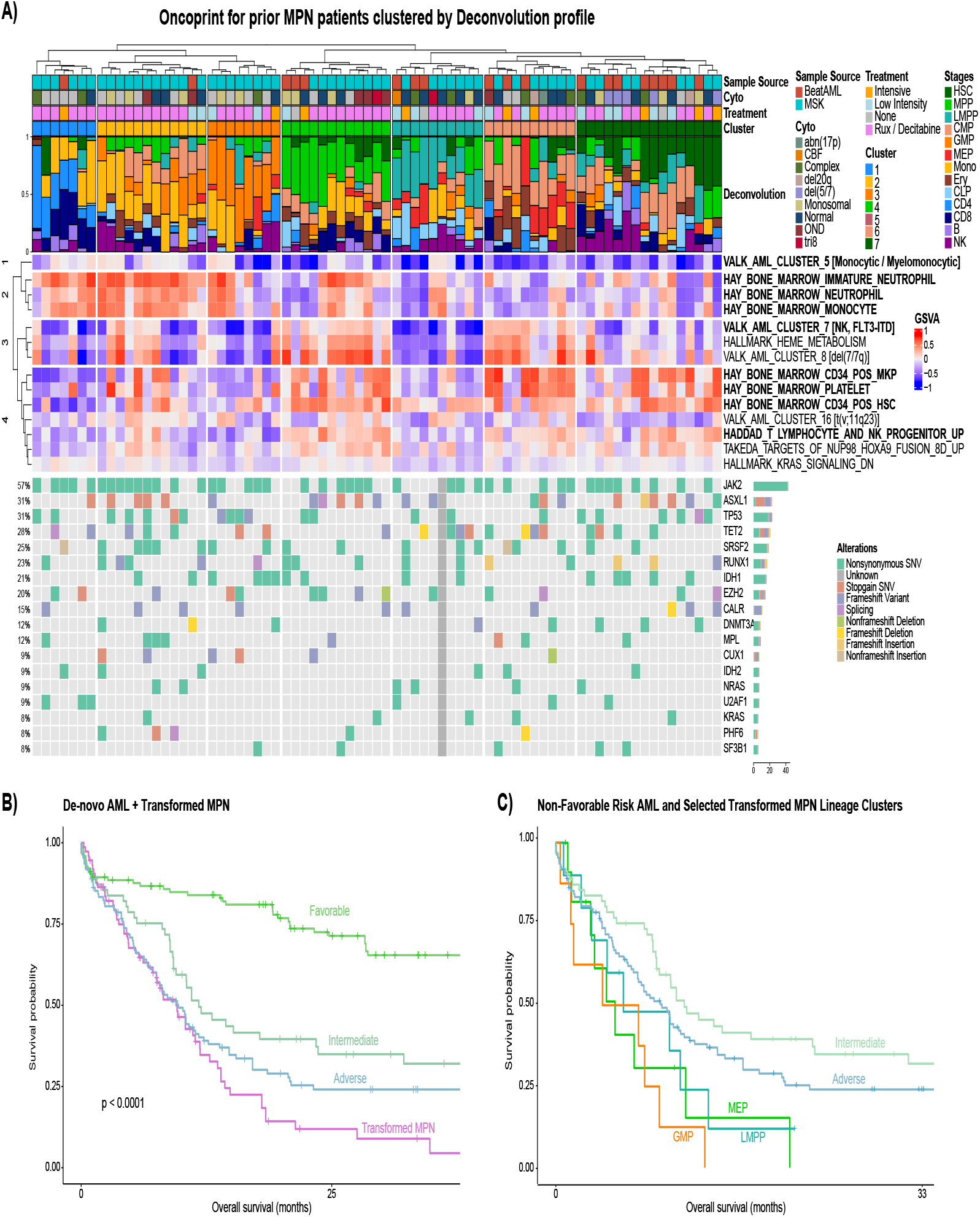
Analysis of transformed MPN. A) Unsupervised analysis of transformed MPN based on lineage composition. Unsupervised hierarchical clustering identified seven distinct clusters corresponding to T cell, monocyte, GMP, MPP, LMPP, CMP, and HSC predominant disease. Cytogenetics, treatment characteristics, Gene set analysis by GSVA, and high frequency molecular features are annotated. B) Survival analysis of transformed MPNs compared to Good, Intermediate, and Poor risk AML. C) Kaplan-Meier curve of overall survival by selected transformed MPN lineage clusters, with intermediate- and adverse-risk AML from the Beat AML cohort shown for comparison.

To determine the impact of lineage character on clinical course, we performed a survival analysis comparing transformed MPN lineage clusters to intermediate- and adverse-risk AML (**Figure 5B, C, S3**). We found significantly worse OS among patients in three transformed MPN clusters: cluster 2 (MPP enriched, p < 0.01), cluster 5 (LMPP enriched, p = 0.038), and cluster 7 (GMP enriched, p < 0.01). We used a Kaplan-Meier estimator to visualize the survival curves for these clusters, with intermediate- and adverse-risk AML plotted for comparison. Here we see that transformed MPN has a median OS that is similar to or worse than patients with adverse risk AML. In addition, we also compared clinical outcomes between all transformed MPN cases and AML. Compared to intermediate risk AML, transformed MPN had significantly worse survival probabilities (HR: 1.79 95% CI: 1.18, 2.71, p = 0.0057), with a median OS that was similar to or worse than adverse-risk AML.

## Discussion

To our knowledge, this is the first description of the transcriptional characteristics of MPN-AP/BP, its comparison to de novo AML, and the first paired comparison of transcriptional and lineage characteristics for patients with chronic-phase MPNs that subsequently transformed. Our study provides valuable insights into the transcriptomic landscape of transformed MPNs (including both accelerated- and blast-phase disease), and builds on prior insights derived from de novo AML^26–30^. By leveraging an integrated analysis with the BeatAML cohort, we are able to draw novel contrasts to de novo AML, including distinct characteristics and quantification of poor outcomes in patients with high-risk transformed MPNs.

We find that MPN-AP/BP is largely a transcriptionally distinct entity from de novo AML, with distinct gene set enrichments. MPN-AP/BP patients tended to have enrichment in stem cell and less differentiated hematopoietic gene sets, whereas de novo AML demonstrated upregulation in later stages of hematopoiesis. Our analysis also revealed that MIR29B1 was substantially upregulated in MPN AP/BP, while the proto-oncogene CT45A1 showed substantially decreased expression. The upregulation of MIR29B1 has been implicated in fibrotic diseases, but its upregulation has not been previously associated with the residual sequelae of MF antecedent to transformation^31–34^. Overexpression of CT45A1 has been implicated in increasing cell stemness in multiple malignancies in cooperation with other genes, and may represent a pathway to malignancy that distinguishes AML from transformed MPNs ^35,36^.

De novo AML and tAML occur as a common manifestation of disparate cellular perturbations, and the same appears true for transformed MPNs^11,37–39^. While MPN-AP/BP does not appear to have a singularly defining set of cytogenetic or molecular features, we found that it could generally be distinguished from AML based on transcriptomic characteristics. Furthermore, transcriptome-derived lineage composition can also define specific subsets of patients with MPN AP/BP including those with HSC, LMPP, MPP, CMP, monocyte, or GMP predominant disease character. While therapies targeting the genomic alterations of MPN AP/BP are under development, we provide a new lens through which the development of novel therapies may proceed – with a focus on specific lineage characteristics^22,40,41^.

Our paired samples allowed to us to garner novel insights from our unique dataset. We observed that canonical pathways associated with MPN pathogenesis, such as IL2 / STAT5 signaling, IL6 / JAK / STAT3 signaling, and NUP98 / HOXA9 fusions, were enriched in the chronic-phase MPN but no longer in MPN AP/BP^42–46^. This implies that, as the MPN transforms, a shift occurs in its transcriptional programming to more primitive of hematopoiesis. This may be due to transformation of the cells that are producing the chronic phase MPN itself, or the secondary induction of an altered bone marrow microenvironment, creating a niche hospitable to the development of more proliferative myeloid clones. Regardless of the specific mechanism, the relative downregulation of these pathways implies that JAK2 inhibition at this phase of the disease will be less effective.

Prior studies have shown that traditional AML therapies including anthracycline-based treatment are less effective in transformed MPNs^24,25^. Our work provides novel insights into the pathophysiology underlying this phenotype, including the enrichment of a doxorubicin resistance gene set among transformed MPNs. Further studies are needed to better understand how this therapeutic resistance may be overcome, either with the addition of targeted agents to anthracycline-containing regimens, avoidance of anthracycline-based therapy (e.g. in favor of high-dose cytarabine or azacytidine +/-venetoclax), or the development of alternative agents entirely. The identification of common lineage features in transformed MPNs despite diverse cytogenetic and molecular features may provide unifying biological insights to help direct novel drug development.

## Data Availability

All data produced in the present study are available upon reasonable request to the authors

https://www.ncbi.nlm.nih.gov/geo/query/acc.cgi?acc=GSE210253

