## Supplementary material for "MPN Transformation Is Characterized By Heterogeneous Shifts In Lineage Character Resulting In Both HSC-Like And More Differentiated Lineage Signatures": NA

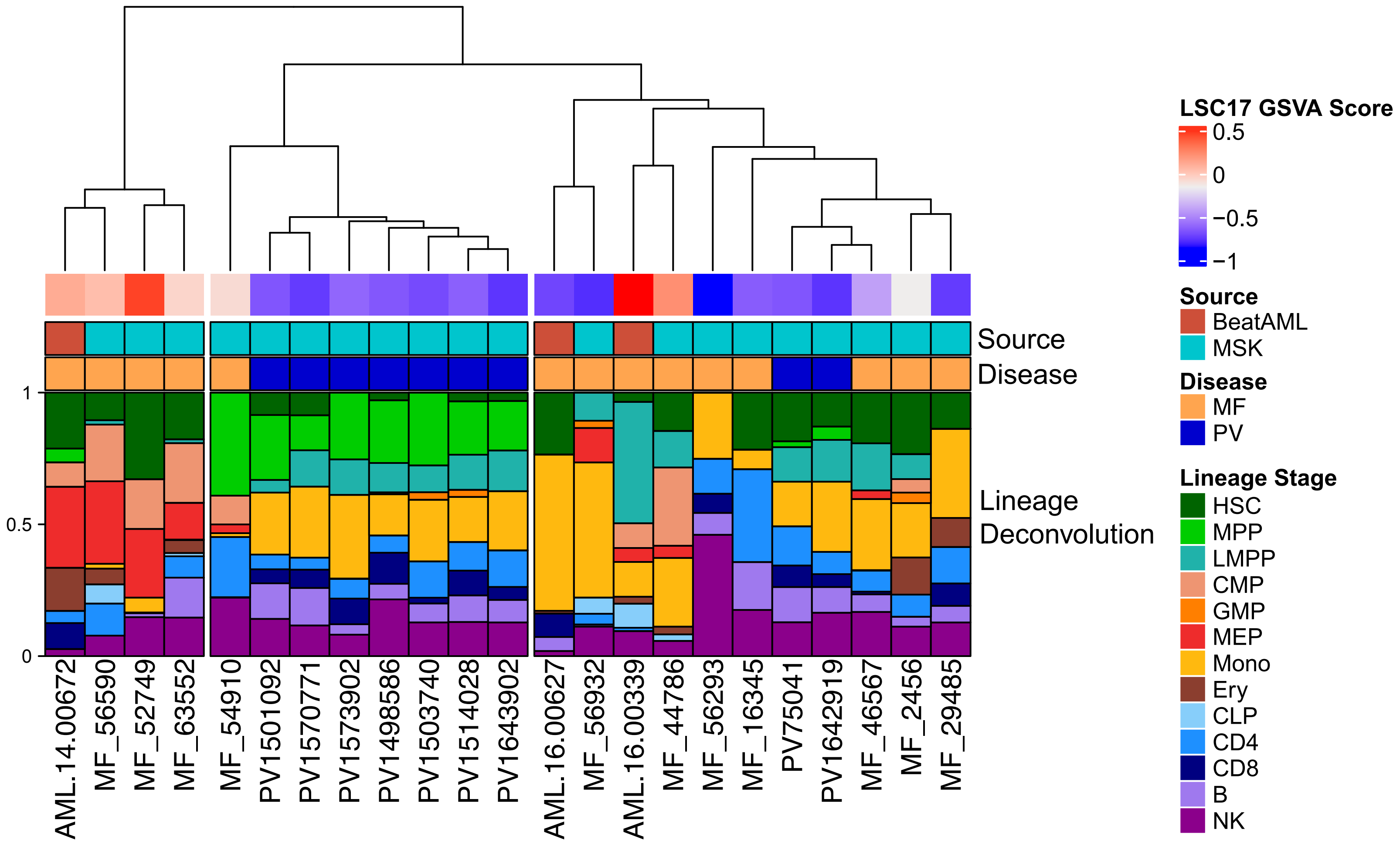


**Figure S1.** *Lineage based analysis of PV and MF transcriptomes.* Lineage deconvolution analysis using our standard bulk RNA-seq pipeline was performed on the subset of PV and MF samples in the combined MSK and BeatAML cohort. Sample source and disease type are shown as annotation labels. In addition, GSVA analysis was performed using the LSC-17 gene signature. Hierarchical clustering was performed based on the resulting deconvolution output using Ward's method (Ward.D2) applied to euclidean distance. The resulting dendrogram was split into 3 clusters by tree height.

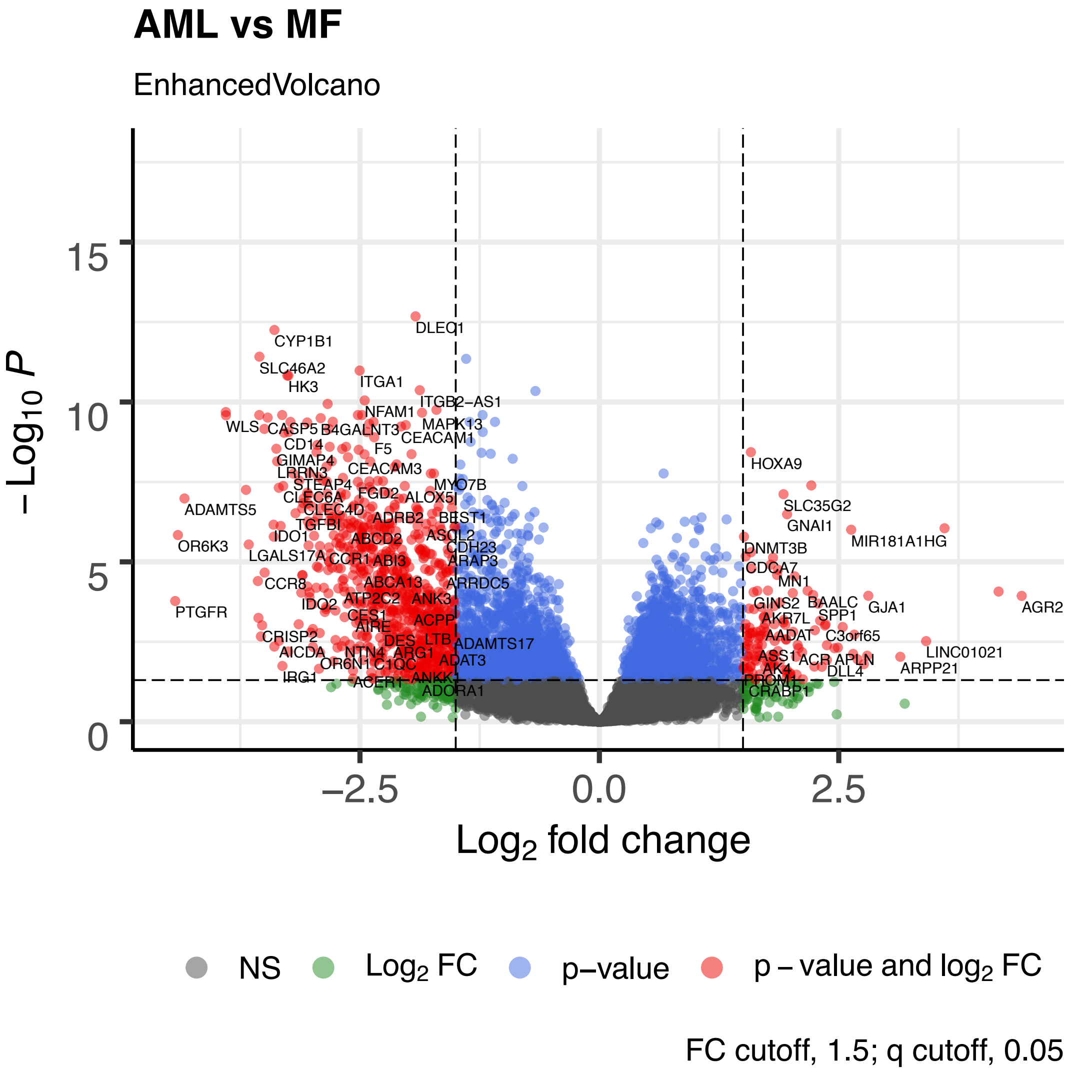


**Figure S2.** *Volcano plot of RNA-seq analysis of MF samples pre and post transformation to AML.* This plot shows the result of DESeq2 based RNA-seq analysis comparing transformed MPN cases (‘AML’) to samples gathered from the same patients prior to transformation. The analysis utilized a model statement incorporating both disease stage and sample identity to adjust for sample specific features. Log fold change of gene expression is plotted against adjusted significance. Genes are colored according to whether the log fold change is above 1.5 (green), q value is less than 0.05 (blue), or both (red). Selected genes with logFC above threshold and a significant q value are printed.

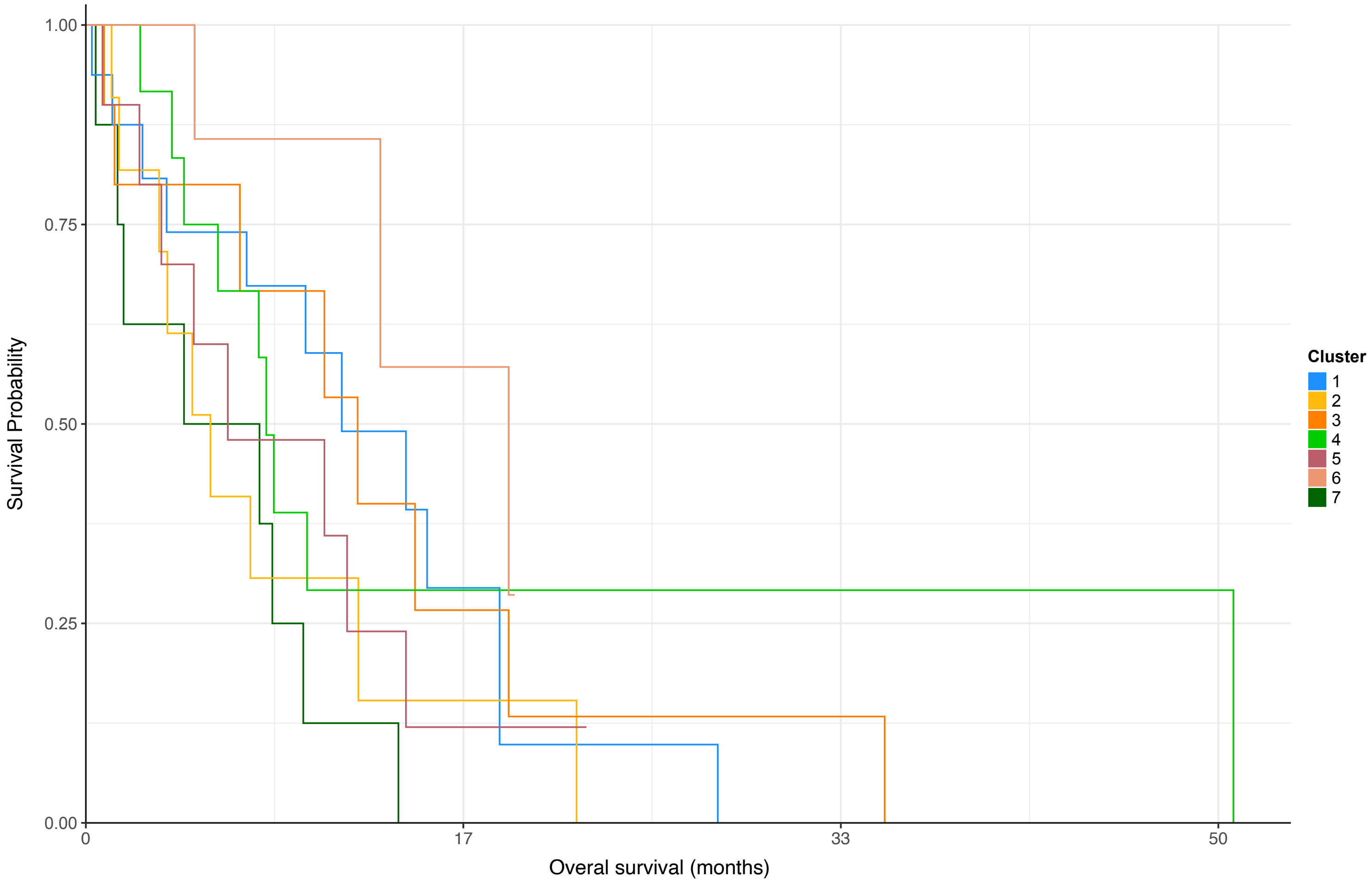


**Figure S3.** *Survival analysis of transformed MPN cases.* Patient groups were determined using the lineage deconvolution-based clustering shown in Figure 5A. Overall survival curves were generated starting at the time of transformation.
